# An inflammation-derived and clinical-based model for ischemic stroke recovery

**DOI:** 10.1101/2024.02.18.24303013

**Authors:** Jiao Luo, You Cai, Peng Xiao, Changchun Cao, Meiling Huang, Xiaohua Zhang, Jie Guo, Yongyang Huo, Qiaoyan Tang, Liuyang Zhao, Jiabang Liu, Yaqi Ma, Mingchao Zhou, Yulong Wang

**Affiliations:** Department of Rehabilitation Medicine, Dapeng New District Nan’ao People’s Hospital, Rehabilitation Branch of the First Affiliated Hospital of Shenzhen University, Shenzhen, China; Department of Rehabilitation, Shenzhen Second People’s Hospital, the First Affiliated Hospital, Shenzhen University School of Medicine, Shenzhen, China; Shenzhen Institute of Translational Medicine, the First Affiliated Hospital of Shenzhen University, Shenzhen Second People’s Hospital, Shenzhen, China; Department of Rehabilitation Medicine, Shandong University of Traditional Chinese Medicine, Jinan, Shandong Province, PR China

**Author notes:** Corresponding author. (Yulong <u>Wang);</u> (Mingchao Zhou). These authors contributed equally, and each has the right to list themselves first in author order on their curriculum vitaes (Jiao Luo, You Cai).

**Keywords:** Ischemic stroke, recovery biomarkers, TIMP1, LGALS3, neuroinflammation

## Abstract

**Background:** Neuroinflammatory responses reflecting disease progression are believed to be closely associated with the severity of prognosis in post-stroke.

**Purpose:** This study developed a combined predicted model of inflammation-derived biomarkers and clinical-based indicators using machine learning algorithms for differentiation of the functional outcome in patients with subacute ischemic stroke.

**Methods:** Clinical blood samples and patient data from individuals with subacute ischemic stroke were collected at admission. Based on activities of daily living assessments followed by a 3-month recovery, patients were categorized into two groups: those with little effective recovery (LE) and those with obvious effective recovery (OE). Serum samples underwent proteomic testing for initial candidates. Subsequently, multidimensional validation of candidates in models of ischemia-reperfusion at protein and mRNA levels was performed. *T*-test, Receiver Operating Characteristic (ROC), and LASSO analysis in an additional cohort were performed to confirm the clinical variables and candidate biomarkers in the discriminatory sensitivity and specificity between the LE and OE groups. Finally, models were developed based on candidates in the training dataset and predicted stroke recovery outcomes in another new dataset using ten standard two-categorical variable algorithms in machine learning.

**Results:** We identified higher tissue inhibitor metalloproteinase-1 (TIMP1) and LGALS3 levels were positively correlated with the severity of prognosis after ischemic stroke rehabilitation. TIMP1 (AUC=0.904, 0.873) and LGALS3 (AUC=0.995, 0.794) were confirmed to address superior sensitivity and specificity in distinguishing ischemic stroke from healthy control and LE group from OE group. The TIMP1 and Lgals3 expression exhibited an evident increase in microglia following ischemia-reperfusion. In addition, inflammation-derived biomarkers (TIMP1, LGALS3) coupled with clinical-based indicators (HGB, LDL-c, UA) were built in a combined model with random forest to differentiate OE from LE in 3-month follow-up with high accuracy (AUC = 0.8).

**Conclusion:** Our findings provided evidence supporting the critical prognostic potential and risk prediction of inflammation-derived biomarkers after ischemic stroke rehabilitation in complementary to current clinical-based parameters.

## 1. Introduction

Ischemic stroke, which accounts for 87% of all strokes, is a leading cause of death and long-term disability worldwide. Approximately 1300 million surviving patients with ischemic stroke suffer from different degrees of disability, which is a substantial burden on the aging population of China^1^. Rapid reperfusion with thrombolysis and/or thrombectomy for ischemic stroke continues to advance^2^, and more attention is focused on how to improve stroke rehabilitation and decline the disability rate. Potential biomarkers of stroke recovery provide knowledge of both therapeutic targets and correlate with the disease severity of rehabilitation. However, there are no biomarkers that have addressed sufficient specificity, sensitivity, and reliability to be applied in the clinical management of patients with stroke, thus highlighting the need for additional study.

Circulating molecules serve as clinically applicable indicators of disease state and progression, reflecting underlying molecular/cellular processes that can be utilized to predict treatment response and stroke recovery. Those molecules may encompass biological markers (blood, genetics), neurological repair markers (electrophysiological activity, tissue remodeling, or neuroinflammation), and clinical test indices (blood routine or urine routine). Multilevel omics have yielded a wealth of promising biomarkers, which serve as clinically relevant indicators for disease state and progression. It has been reported that high-density lipoproteins^3^, phenylacetylglutamine^4^, and serum cytokines^5^ could represent independent prognostic markers and inform on stroke recuperation. The variety of promising predictors regarding which ones possess superior predictive value is also confusing. The emergence of machine learning algorithms may offer non-invasive approaches to further screen for more valuable variables and construct combined models, effortlessly incorporating a vast number of variables. These characteristics make machine learning remarkably efficient and shiny to apply in the medical domain.

The inflammatory response has been implicated in the development of brain ischemic pathology and plays a pivotal role in tissue remodeling and repair during stroke recovery. The acute stage of cerebral ischemic injury and reperfusion, lasting approximately one week, triggers an inflammatory cascade mainly characterized by inflammatory cell infiltration, inflammatory mediators release, blood-brain barrier damage, extracellular matrix degradation, oxidative stress, and excitotoxicity. These factors further contribute to neurovascular unit (NVU) damage and death. In the subacute stage (within six months) after ischemic stroke, microglia play both neurotoxic and protective roles that account for the complexity of the immune response^6^. Ischemic stimulation leads to the alterations of microglia activation and proliferation with M1-liker polarization releasing pro-inflammatory factors such as TNF-α, NOS, CXCL10, MMP9, and M2-liker polarization releasing anti-inflammatory factors such as IL-10, Arginase-1, CD206, IL-4, IL-13, and IL-33^7^. Preclinical studies have demonstrated significant changes in specific genes of different types of cells within the NVU following middle cerebral artery occlusion (MCAO) through single-cell sequencing analysis, among which 60% are microglia-specific genes^8, 9^. Targeting the central immune role of microglia may hold promise as a critical therapeutic approach for regulating neuroimmunity in post-stroke rehabilitation.

In this study, we hypothesized that ischemic stroke induces characteristic molecular changes associated with inflammatory responses and predicts functional recovery from stroke events. Thus, we aimed to develop a combined model of inflammation-derived biomarkers and clinical-based parameters model using machine learning algorithms to predict ischemic stroke rehabilitation.

## 2. Methods

### 2.1 Study design and participants

This study was conducted at the Rehabilitation Department, Shenzhen Second People’s Hospital. All the cases of 52 male patients with ischemic stroke and 20 male healthy controls were collected from June 2020 to June 2022. The recovery effect of the enrolled patients with ischemic stroke in daily activity ability was assessed by the Longshi Scale and Barthel index (BI) after the patients had received conventional rehabilitation for 3 months. Longshi Scale, a supplement for modified Rankin Scale (mRS) and BI, divided disabled people into Bedridden, Domestic, and Community groups according to the person’s daily activity ability and activity range^10^. The group of little effect recovery (LE) defined in a status of from Bedridden (at admission) to Bedridden (3 months after discharge) group and obvious effect (OE) recovery from Bedridden (at admission) to Domestic group (3 months after discharge) were assessed by the Longshi Scale as previously discribed^10^. Patients with severe systematic or mental diseases, other causes of brain injury, or sharp, blunt direct action on the head caused by organic brain tissue damage were excluded from this study.

In brief, 30 LE patients with poor prognosis and 22 OE patients with good prognosis were qualified, together with 18 healthy controls. Subjects were divided into discovery groups, validation groups, and new test prediction groups, schematically summarized in Figure 1.

**Fig1.**
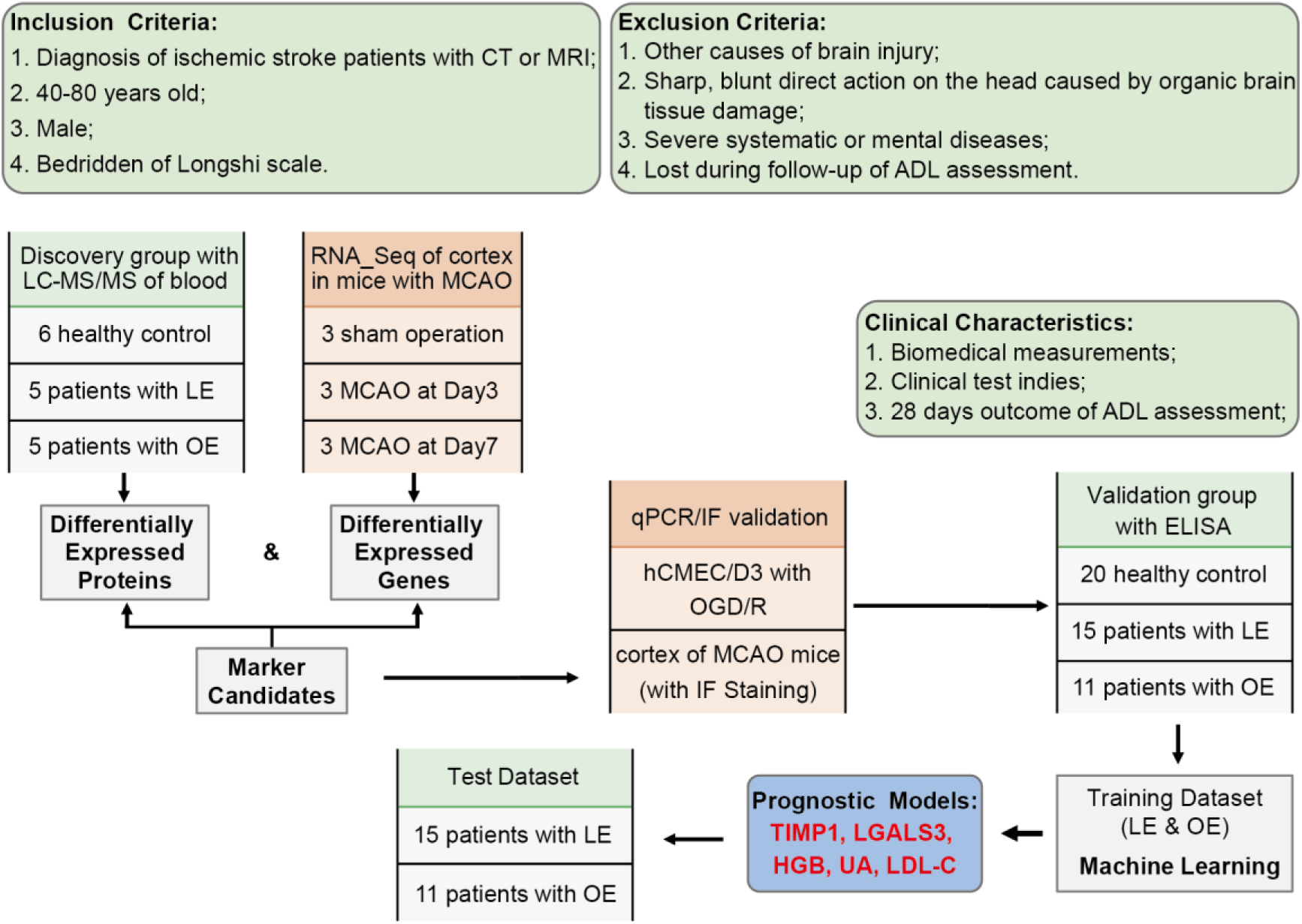
Overall experimental design for a combined model development based on machine learning. Ischemic stroke patients were categorized into LE and OE group based on activities of daily living assessments followed by a 28-day recovery. Discovery group underwent proteomic testing for initial candidates. Multidimensional validation of ischemia-reperfusion at protein and mRNA levels was performed both in vivo and in vitro. ROC and LASSO analysis in an additional cohort to confirm the candidate biomarker and clinical variables in the discriminatory sensitivity and specificity between the LE and OE groups. Candidates were modeled using ten standard machine learning algorithms and then prediction outcomes in another new dataset. **Abbreviations:** CT, Computed Tomography; MRI, magnetic resonance imaging, LC-MS/MS, liquid chromatography-tandem mass spectrometry analysis; LE, little effective recovery; OE, obvious effective recovery; HC, Healthy Control; MCAO, middle cerebral artery occlusion; OGD/R, oxygen-glucose deprivation/ reoxygenation; HGB, hemoglobin; LDL-c, low-density lipoprotein cholesterol; UA, uric acid; ADL, activities of daily living.

### 2.2 Collection of serum samples

The blood samples of patients enrolled were obtained on the second day after hospitalization at the Department of Rehabilitation Medicine. The samples were promptly processed according to the standardized protocol recommended by the HUPO Plasma Proteome Project. Briefly, blood was drawn into plastic K2EDTA tubes (BD), gently inverted manually ten times, and stood at room temperature for 1 hour. Subsequently, the blood samples were centrifuged at 4°C and 1000 × g for 10 minutes, and serum sample aliquots were stored at -80°C until further proteomic analysis or ELISA.

### 2.3 LC-MS/MS and bioinformatic analysis of serum proteomic

The serum samples of a discovery cohort were transported with dry ice to Jingjie PTM BioLab (Hangzhou) Co. Ltd. for standardized proteomic sample pretreatment and four-dimensional label-free quantification of Liquid chromatography-tandem mass spectrometry analysis (LC-MS/MS) as the previous described^11^. After removal of high-abundant proteins using Pierce™ Top 14 Abundant Protein Depletion Spin Columns Kit (Thermo Scientific), trypsin digestion of the proteins was dissolved in solvent A (0.1% formic acid in water) and separated with a gradient solvent B (0.1% formic acid in acetonitrile). The gradient settings were: 4%-6% solvent B in 2 min; 6%-24% solvent B over 68 min; 24%-32% solvent B in 14 min; 80% solvent B in 3 min; then holding at 80% for the last 3 min, all at a constant flow rate of 300 nL/min on a nanoElute ultra high-performance liquid chromatography (UHPLC) system (Bruker Daltonics). The peptides were subjected to a capillary source followed by the timsTOF Pro (Bruker Daltonics) mass spectrometry (1.60 kV electrospray voltage) in parallel accumulation serial fragmentation (PASEF) mode. Precursors and fragments were conducted an MS/MS scan (100 to 1700 m/z) at the TOF detector. Precursors with charge states (0 to 5) were selected for fragmentation, and 10 PASEF-MS/MS scans were acquired per cycle. The dynamic exclusion was set to 30 seconds.

The MS/MS data were retrieved by the Proteome Discoverer (v2.4.1.15). A human database was searched by Homo_sapiens_9606_PR_20201214.fasta (75777 protein sequences). The decoy database antilibrary was used to reduce the false-positive rate (FPR). The FDR was adjusted to <1%, and the minimum score for modified peptides was set to >40. Differentially expressed proteins (DEPs) were considered separately with log2|foldchange| ≥1.0 between the LE, OE, and HC groups. Based on the protein sequence alignment method, the protein domain functions were defined by InterProScan (http://www.ebi.ac.uk/interpro/). Principal Component Analysis (PCA) was performed using the FactoMineR (version 2.4) and factoextra (version 1.0.7) packages in R to reduce dataset dimensionality and visualize relationships among variables and samples.

### 2.4 Total transcriptome sequencing and bioinformatics analysis

The transcriptome sequencing of cerebral ischemic stroke was conducted by reusing a previously published study^12, 13^, in which our co-first author (You Cai. PhD) played a key role. The transcriptome sequencing data has been published and can be found in the Genome Sequence Archive with accession numbers CRA001143 and CRA001432. All expressed and differentially expressed genes (DEGs) were determined based on |log2FoldChang|>0 and a *p* < 0.05. Overlaps of the DEPs of plasma proteomic and DEGs in mice with middle cerebral artery occlusion (MCAO) at day 3 and day 7 were obtained with a Venn diagram. Protein-protein interaction of the overlaps was visualized and analyzed with Cytoscape. Gene Ontology (GO) annotation analysis was executed with the clusterProfiler R package (version 4.2.2) applying parameters such as ’pAdjustMethod = BH, p-value < 0.05, and simplify cutoff = 0.5’. At the same time, default settings were maintained for the remaining parameters. ROC analysis was performed using the pROC R package (version 1.18.0).

### 2.5 MCAO modeling and 2,3,5-Triphenyl Tetrazolium Chloride (TTC) Staining

Male C57BL/6J mice (6-8weeks) were purchased from Guangdong Weitong Lihua Experimental Animal Technology Co., Ltd. Guangzhou, China, and housed under specific pathogen-free conditions with 22°C ± 2°C and a 12-light/dark cycle. All procedures were approved by the Experimental Animal Welfare and Ethics Committee of Shenzhen Institute of Translational Medicine, Shenzhen Second People’s Hospital.

A silicon-coated tip (Jialing Biotech, China) and C57BL/6 male mice (25-30 g) at 12 weeks were applied to establish a mouse model of ischemic stroke, according to a previous study^13^. In brief, mice were anesthetized with 1-2% isoflurane and maintained using Small Animal Anesthesia Machine R500 (RWD Life Science, China). During MCA occlusion surgery, a silicon-coated tip was introduced from the proximal end of the external carotid artery to the distal end of the external carotid artery. The tip was kept in place for 1 hour (occlusion), then removed and sutured (refill). The sham operation mice were treated with similar surgery, except the tip was not inserted. After mice regained full consciousness, neurological severity scores were assessed using a five-point scale according to a previous study^14^.

TTC stain was performed to evaluate the ischemic areas of the MCAO model. Mice were anesthesia with intraperitoneal injection of 1% pentobarbital sodium 24 hours after the MCAO surgery. Then, the intact brain was isolated, rapidly frozen at -20℃ for 10 min, placed in a mouse meningeal capsule, and prepared for seven coronal (1mm) per mouse. Subsequently, brain sections were stained with 2% TTC solution (Cat. T8877, Sigma-Aldrich) at 37℃ away from light for 15 min.

### 2.6 Cell culture and Oxygen-glucose deprivation/ reoxygenation (OGD/R)

The human brain microvascular endothelial cell line (hCMEC/D3) was purchased from Sepkon (iCell-h070). It was cultured normally in Endothelial Cell Medium (ECM) (Cat. 1001, ScienCell) supplemented with Endothelial Cell Growth Supplement (ECGS, Cat #1052, ScienCell) and 5% fetal bovine serum (FBS). The mouse microglial cell line (BV2) was presented from Northeastern University and cultured in DMEM/HIGH glucose medium containing 10% FBS. In control, the two cell lines were cultured in a conventional medium and placed in an incubator with an atmosphere of 5% CO2/95% air.

For OGD/R, the cells were seeded into a 6-well plate at 3 × 10^5^ /well and grew approximately 70-80% confluency; the medium was replaced by MEM medium without glucose (Cat#A1443001, Gibco). Then, cells were placed into a humidified 37℃ incubator with a gas mixture of 1% O2, 5% CO2, and 94% N2 at the control of ProOx C21 (Biospherix, USA). After 6 hr exposure to hCMEC/D3 and 4 hr exposure to BV2, the cells were cultured with reperfusion of oxygen and nutrients.

### 2.7 Real-time PCR Analysis

The cortex tissues or cells were collected, and total RNA was extracted with TRIzol reagent (Cat.15596026, Invitrogen) following the manufacturer’s instructions. One µg of total RNA from each sample was reverse transcribed to cDNA with RevertAid RT Reverse Transcription Kit (Cat.K1691, Thermo Scientific™). qPCR was performed with SYBR Green Mix (Cat.QPK-201, Toyobo). Results were collected with Bio-Rad CFX Connect Real-Time system and presented as linearized values normalized against β-actin in triplicate. Suzhou Hongxun Biotechnology synthesized the primers (Table S1).

### 2.8 Immunofluorescence of brain tissue

Mice subjected to MCAO operation on day 7 were perfused with 0.9% saline flush and 4% paraformaldehyde (PFA) after cardiac blood collection. Intact brain tissue was isolated and fixed with 4% PFA for 24 h, then dehydrated with 20% sucrose and 30 % sucrose. Brain tissue was placed in O.C.T. media and froze at -20℃. Contiguous coronal sections taken across the hippocampus were performed for double-immunofluorescence using a rabbit polyclonal antibody for Iba1 (WFD6884, 1:500) and Timp1(sc-21734,1:100) or Galectin3 (sc-32790, 1:100) mouse monoclonal antibody. The immunofluorescence in brain slices from the cortex and the hippocampus was visualized by confocal microscopy (KEYENCE BZ-X, Japan).

### 2.9 Validation study of targeted biomarkers with ELISA analysis

For the validation study, 100 μL serum was separated from the collected sample. The levels of targeted proteins in serum were detected according to the manufacturer’s instruction by corresponding kits of enzyme-linked immunosorbent assays, such as TIMP1 (RDR-TIMP1-Hu, Reddot Biotech) and TGFB1 (EK981, MULTI SCIENCES) LGALS3 (EK1126, MULTI SCIENCES). Serum samples were diluted by a factor of 10.

### 2.10 Machine learning in constructing a predictive model integrating biomolecules and clinical indicators

The Lasso algorithm was employed for the initial screening of predictor variables for machine learning. Variables identified by Lasso underwent further screening, with those exhibiting high covariance excluded one by one using logistic regression to identify features with p-values < 0.05. A total of 10 different machine learning classification algorithms for distinguishing dichotomous variables were utilized, including Naive Bayesian Classifier (nb), Decision Tree Algorithm (C5.0, AdaBag, and Random Forest), Support Vector Machine (svmRatial, svmPloy, and svmLinear), Logistic Regression (glmnet), K-Nearest Neighbor (kknn), Artificial Neural and Network (nnet). These algorithms were integrated within the caret (version 6.0.92) (74) R package.

### 2.11 Statistical Analysis

The statistical analysis is detailed in the Data Processing and Analysis sections. All other clinical and laboratory data statistical analyses were conducted using GraphPad Prism (v8.0). Results of normally distributed parameters are expressed as mean ± SEM. Data were compared by a two-tailed unpaired Student’s *t*-test or one-way analysis of variance for multiple comparisons. When necessary, experimenters were blinded to group allocation prior to data acquisition. Significance levels of p-values are provided in the corresponding figure legends. In general, differences were considered statistically significant at *p-values* < 0.05 in all cases. “*” represents *p* < 0.05, “**” represents *p* < 0.01, “***” represents *p* < 0.001, and “****” for *p-values* < 0.0001.

## 3. Result

### 3.1 Inflammation-derived biomarkers robust increasing after ischemia-reperfusion injury

To obtain reliable candidates for differentiation severity of functional outcome, serum proteomics of patients with ischemic stroke and RNA sequence of MCAO animals were applied and integrated to screen differential molecules in this study as schematically summarized in Figure 2A. 5 patients with LE, 5 patients with OE, and 6 HC were selected as listed in Table 1 and Table S2. The serum was collected from the enrolled and subjected to tryptic digest with high abundance removal, followed by proteomics with LC-MS/MS analysis. Based on a shotgun proteomics approach, 1533 proteins were identified from 9422 unique peptides with a maximum false positive rate (FPR)<1%, among which 948 proteins (Figure 2B) at a quantifiable level were used for principal component analysis (PCA) (Figure 2C). The detailed annotation information of 1533 proteins were shown in Table S2. 194 DEPs varied in the LE compared to the HC and 174 DEPs in the OE compared to the HC according to *p*-value < 0.05 (Table S3). On the other hand, the RNA sequence of the ischemic core of the cortex in mice with MCAO at Day 3 and Day 7 was analyzed, and DEGs were taken *p*-value < 0.05 as the standard (Table S4). We found 27 overlaps (Figure 2D) between the DEPs and DEGs. According to the standard of more than 5 interactions, 8 target molecules of TIMP1, LGALS3, VIM, TGFB1, MYH9, CSF1R, PTPRC, and GSN were noticed following protein interaction network analysis (Figure 2E). Next, in the biological process classification of 27 overlaps, terms directly related to the 8 most important protein enriched in negative regulation of cell adhesion, regulation of cell morphogenesis, leukocyte proliferation, T cell activation, plasma membrane organization, actin cytoskeleton reorganization, tissue remodeling, glial cell differentiation (Figure 2F and Table S5), which indicated that inflammation response was enhanced in the subacute ischemic stroke.

**Fig2.**
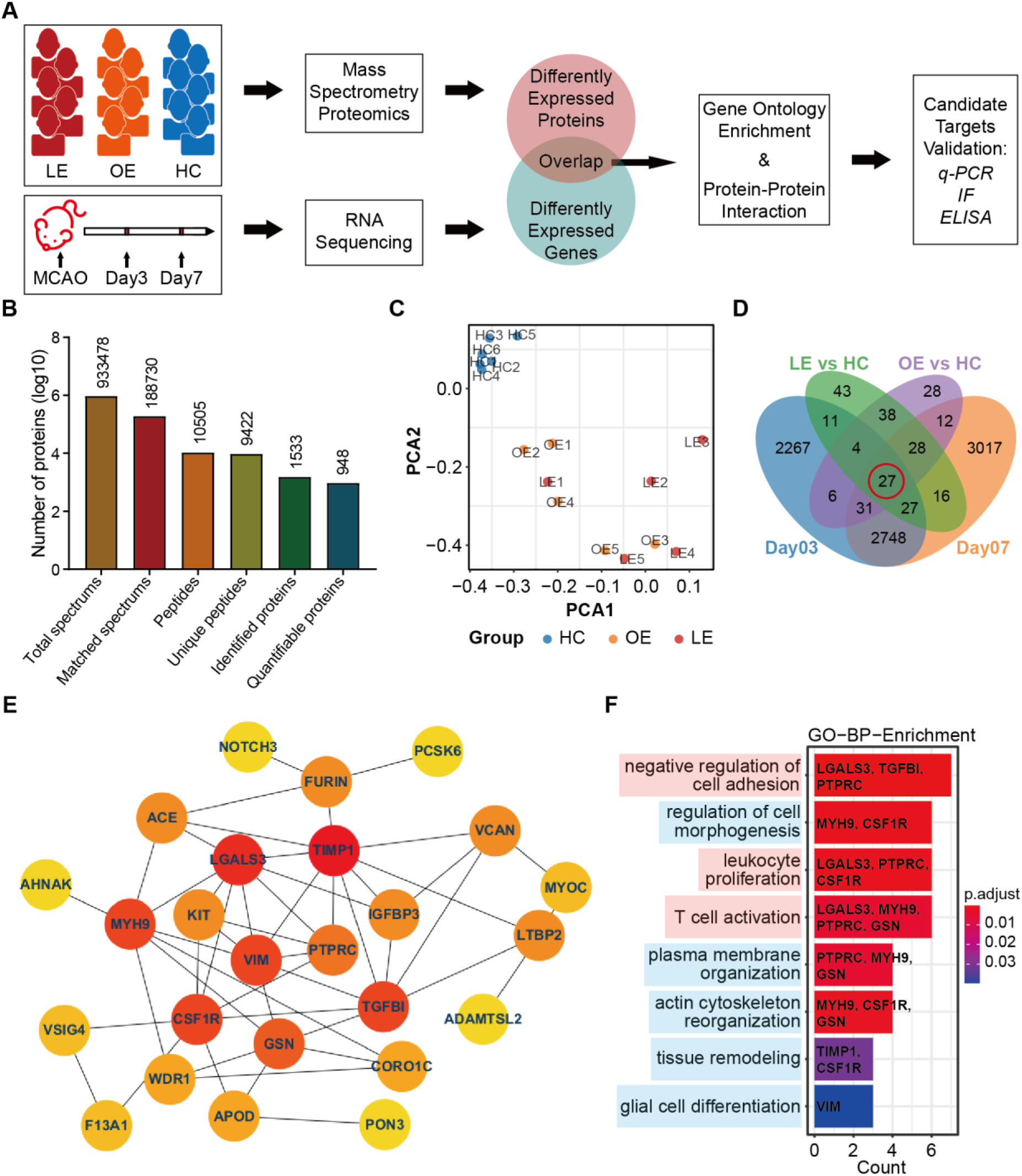
Screening robust target molecules from different omics data of humans and mice after ischemic injury. **A,** Workflow chart of the study. Human serum samples for LC-MS/MS were categorized into three groups based on ADL assessment using the Longshi scale: LE group, representing patients with poor prognosis transitioning from bedridden to bedridden; OE group, representing patients with good prognosis transitioning from bedridden to domestic assessed by Longshi Scale; and healthy controls. Mouse brain samples for RNA sequencing were collected from the operative side of the brain on days 3 and 7 after MCAO, while the contralateral side of the brain served as a control. The differentially expressed proteins (DEPs) obtained through protein profiling and differentially expressed genes (DEGs) obtained through RNA sequencing were overlapped. GO functional analysis and protein network interaction analysis were performed on the intersecting genes. Candidate targets were validated with qPCR, WB and ELISA analyses at both the RNA and protein levels. **B,** Protein information detected by mass spectrometry proteomics in the LE (n=5), OE (n=5), and HC (n=5) groups. **C,** PCA plots revealed the separation of samples in LC-MS/MS. **D,** Venn diagram of differentially expressed proteins obtained by protein profiling and the differentially expressed genes obtained by RNA sequencing, 27 molecules were differentially expressed in all 4 groups. **E,** The protein-protein interaction network diagram displayed by cytoscape. Different colors represent the order of protein importance, the redder the color, the more important it is. **F,** The 27 proteins in the intersection set in D were subjected to GO functional enrichment, and the de-dundantitems terms directly related to the 8 most important proteins in E were filtered from the significantly Biological Processes terms. Shown are the top 8 terms with “p.adjust” ranking. The background colors of the labels were artificially divided into two groups according to function, with the pink group being related to immunity and the blue group being related to histomorphology. **Abbreviations:** PCA, Principal Component Analysis; TGFB1, Transforming Growth Factor beta 1; PTPRC, Protein Tyrosine Phosphatase Receptor Type C; VIM, Vimentin; MYH9, Myosin Heavy Chain 9; CSF1R, Colony stimulating factor 1 receptor; GSN, Gelsolin; GO, Gene Ontology; BP, Biological Processes.

**Table 1.** ADL assessment of enrolled subjects for LC-MS/MS.

| Characteristic | Health Controls<br>(HC) | Ischemic Stroke (IS) |  |
| --- | --- | --- | --- |
|  |  | Obvious effects (OE) | Little effects (LE) |
| Gender (male) | 6 | 5 | 5 |
| Age (year) | 56.50 ± 2.655 | 67.40 ± 4.445 | 67.20 ± 2.478 |
| Diagnosis | Normal | Ischemic stroke | Ischemic stroke |
| ADL assessment of Barthel Index at admission | NA | 34.00 ± 3.674 | 21.00 ± 2.739 |
| (at discharge) |  | (42.00 ± 5.831) | (22.00 ± 2.550) |
| ADL assessment of Longshi Scale at admission | NA | Bedridden group | Bedridden group |
| (at discharge) |  | (Domestic group) | (Bedridden group) |

Furthermore, the 8 target molecules were validated with qPCR in vitro and vivo (Table S6), flow chart shown in Figure 3A. Morphological observation showed that hCMEC/D3 atrophied compared with the control group after OGD 6 hours, and the proliferation was inhibited, followed by 24 hours of reoxygenation (Figure 3B). The qPCR results showed a significant increase of *TIMP1, MYH9, TGFB1, VIM,* and *LGALS3* mRNA (Figure 3C) and no significant change of *CSF1R* and *GSN* mRNA (Figure S1A) in hCMEC/D3 with OGD/R treatment. Meanwhile, the MCAO mice were established and confirmed with TTC straining (Figure 3D). mRNA change in the ischemic core of the cortex isolated from mice with MCAO on day 3 and day 7 was detected by qPCR. Data showed only the changes of *Timp1*, *Tgfb1*, and *Lgals3* mRNA (Figure 3E) were consistent as in the hCMEC/D3 cells and apparent up-regulation, *Csf1r*, *Gsn*, *Myh9, Ptprc*, and *Vim* mRNA without significant expression were shown in Figure S1B.

**Fig3.**
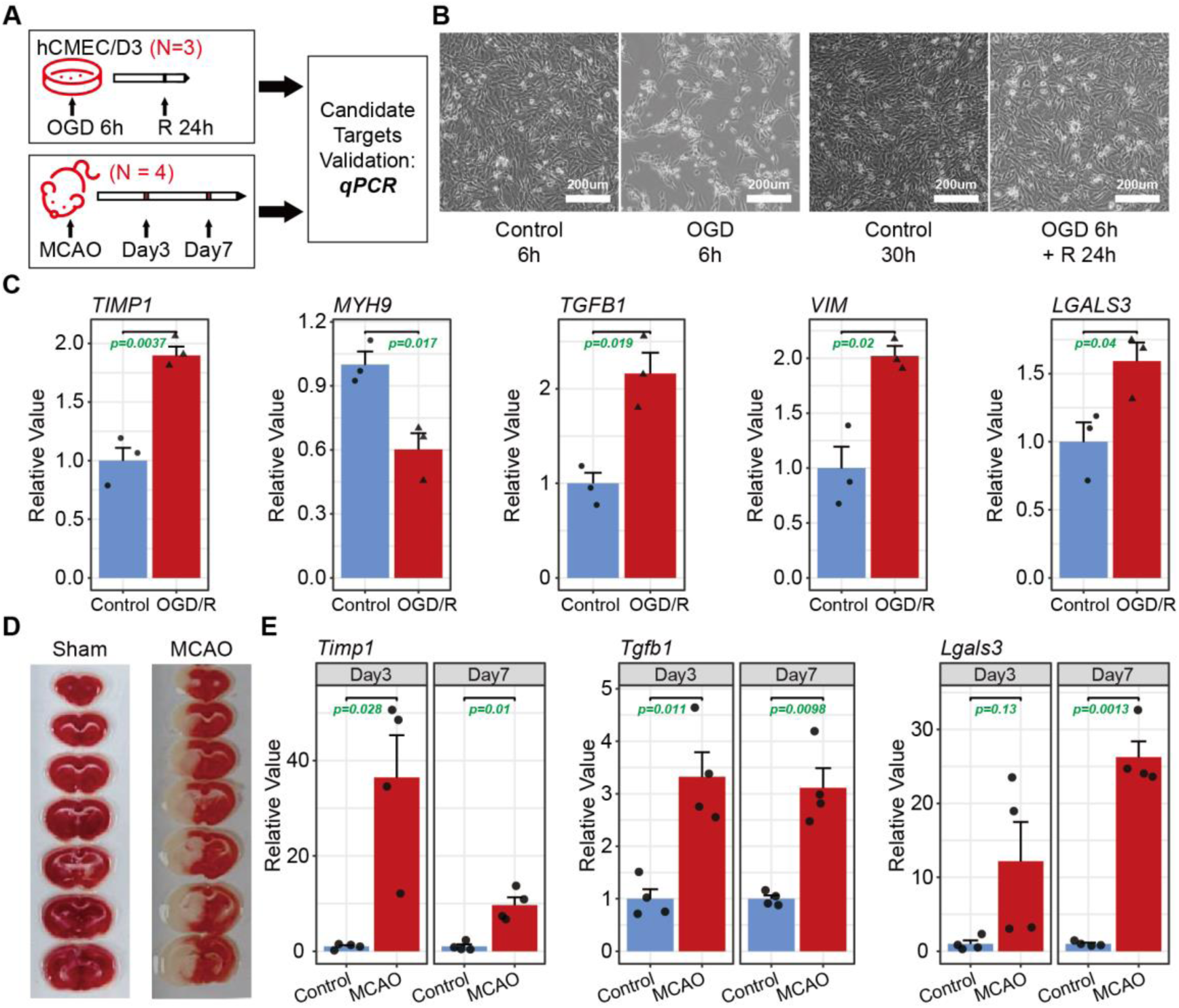
Validation of the candidate targets at the RNA levels in vitro and in vivo. **A,** Work flow chart of study. The hCMEC/D3 cells are human brain microvascular endothelial cell line that were OGD-treated for 6 hr, reoxygenation for 24 hr, and then the samples were collected in 3 biological replicates per group. The model of mice with MCAO was followed by taking brain tissue on days 3 and 7, 4 biological replicates per group. **B,** Bright field images of the hCMEC/D3 cells during OGD/R modeling indicating successful modeling. **C,** Histograms of the q-PCR expression levels of the 8 target molecules in hCMEC/D3 cells with OGD/R. The histogram displays the qPCR expression levels of the 8 target molecules in hCMEC/D3 cells. The statistically significant molecules, with differences determined by *p*-value from smallest to largest, were arranged from left to right. Supplementary Figure S1A showed the molecules without significant differences. Each data point represents a biological sample. **D,** TTC staining images of successful MCAO modeling. **E,** Histograms of the qPCR expression levels of the 8 target molecules of the brain tissue in mice with MCAO. 3 molecules with significant difference presented as in C and Other molecules were shown in Supplementary Fig. 1 B. Each point represents a biological sample.

TIMP1 and LGALS3, recently served as novel inflammatory factors, have a novel role in immune regulation, inspiring attention and exploration. Hence, we detected the changes of Timp1 and Lgals3 expression in brain microglia by IF staining. In an immunofluorescence brain of mice with MCAO on 7 days, we found that the expressions of Timp1 and Lgals3 increased robustly after mice with ischemia-reperfusion injury, especially in the ischemic core of the cortex (Figure 4A and 4F). Iba1 is one of the classic markers of microglia activation. A remarkable increase was observed in Iba1 expression, which suggested that the microglia activation surged in ischemic brain tissue, including the ischemic penumbra cortex, hippocampus, striatum, and hypothalamus. The amplifying image for the corresponding area in Figure 4A showed that most TIMP1 and Iba1 were colocalized in the ischemic cortex (Figure 4B-C) and hippocampus (Figure 4D-E), as well as the colocalization of Lgals3 and Iba1 (Figure 4G-J). Therefore, multidimensional verification of candidate expression drew attention to TIMP1, LGALS3, and TGFB1, closely related to inflammatory response and prognosis.

**Fig4.**
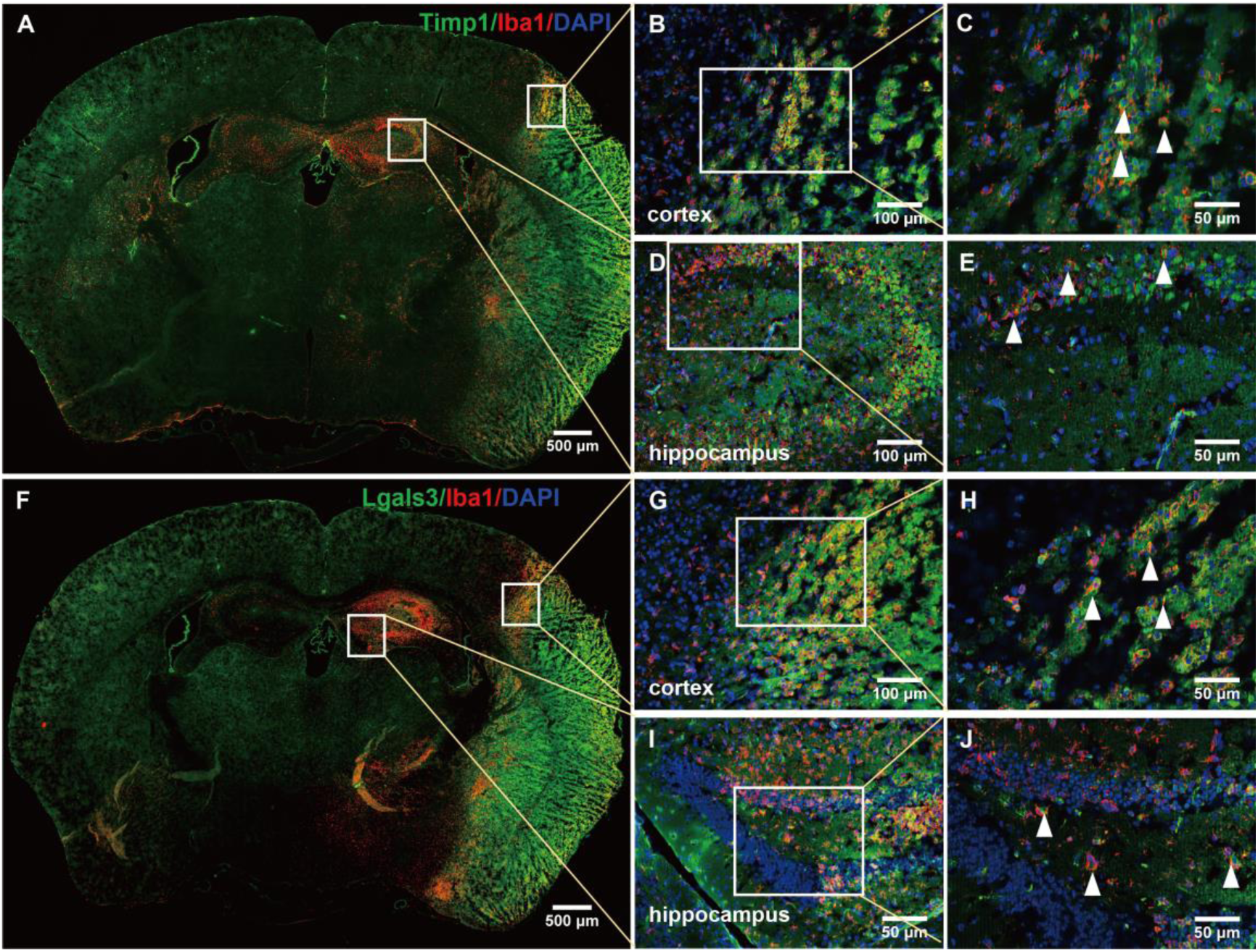
IF staining of candidates in brain sections of mice with MCAO. **A,** Immunofluorescence staining with anti-Timp1 (green) and anti-Iba1 in brain sections of mice with MCAO on day 7; **B-C,** Staining magnification of the ischemic penumbra cortex; **D-E,** Staining magnification of the ischemic hippocampus; **F,** Immunofluorescence staining with anti-Lgals3 (green) and anti-Iba1 in brain sections of mice with MCAO on day 7; **G-H,** Staining magnification of the ischemic penumbra cortex; **I-J,** Staining magnification of the ischemic hippocampus. Nucleus were dyed with DAPI staining (blue). The white triangle refered to the colocalization expression of targets and Iba1.

### 3.2 Biomarker validation by ELISA and ROC analysis

Furthermore, we conducted an ELISA analysis to investigate the serum protein level of TIMP1, LGSAS3, and TGFB1 in an enlarged clinical sample. This analysis included 15 patients with LE and 11 patients with OE who suffered from ischemic stroke events within 3 months and 18 healthy controls. Detailed demographic information and primary ADL assessments for these two outcome groups are presented in Table S7 and Figure S2A-B. Age analysis revealed no significant difference between the healthy controls and stroke patients, while the mean age of the OE group (mean=63.27 ± 2.711) was lower than the LE group (mean=69.80 ± 2.322) (Figure 5A). ELISA data (Table S7) showed that the TIMP1 and LGALS3 levels significantly increased after ischemic stroke; additionally, higher serum TIMP1 and LGALS3 levels during the subacute phase of ischemic stroke were associated with greater severity in ADL activities (Figure 5B-C). However, no difference was observed in TGFB1 levels among the groups (Figure 5D). The lack of association between circulating TGFB1 levels and function outcomes of stroke with ADL assessment is hardly controversial^15^; TGFB1 change was not considered in the following study.

**Fig5.**
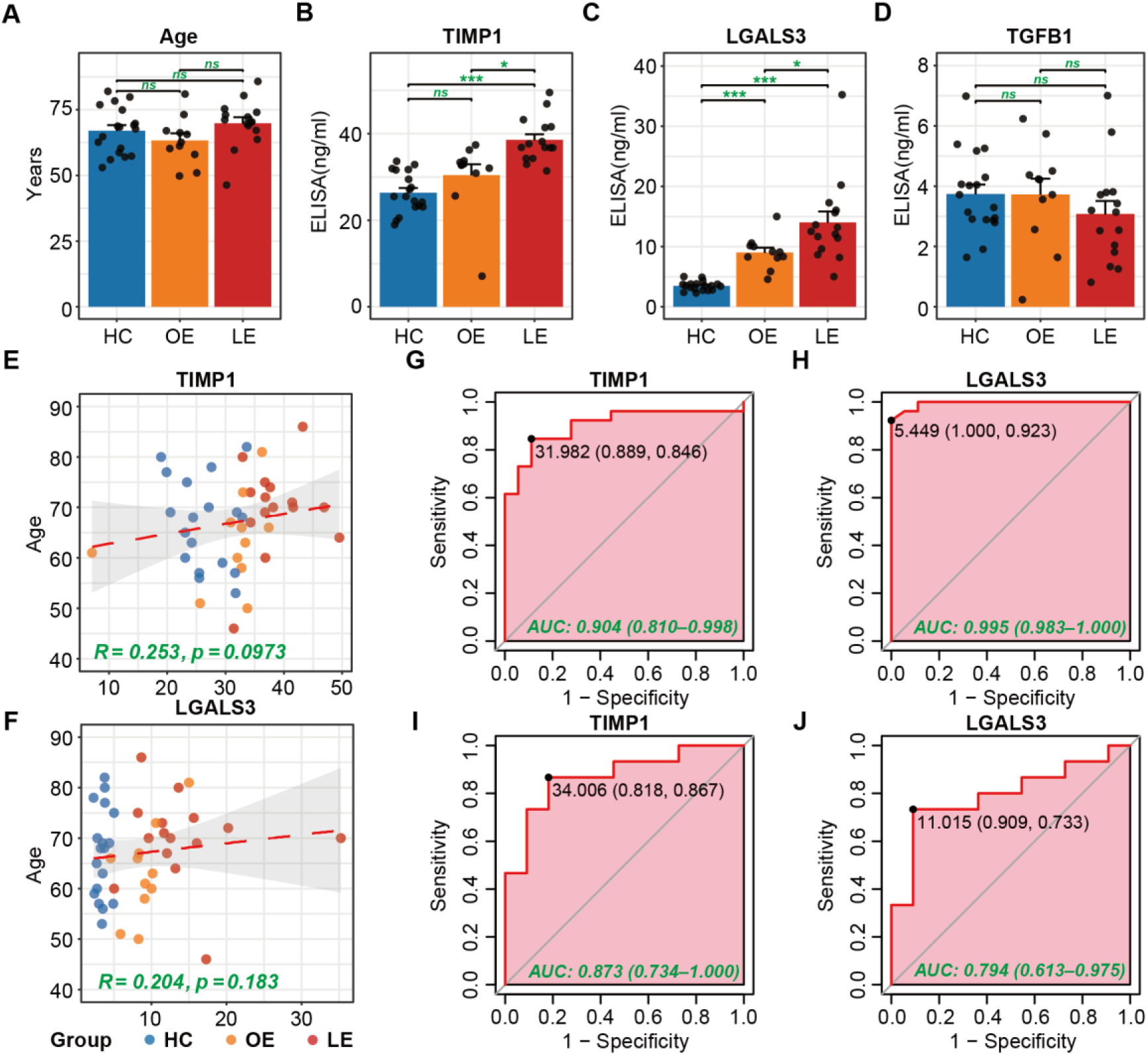
Validation and ROC analysis of target molecules at the protein level in ischemic stroke patients. **A,** Age distribution statistics of recruited clinical subjects of the LE (n=15), OE (n=11), and HC (n=20). Each point represents an individual. **B-D,** Histograms of the ELISA expression levels of the 3 target molecules validated at RNA level. The data were analyzed using one-way ANOVA analysis. “*” represents *p* < 0.05, “**” represents *p* < 0.01, “***” represents *p* < 0.001. Each point represents an individual. **E-F,** Correlation scatter plots of TIMP1 and LGALS3 with age. The red dashed lines are the trend lines, with the Pearson’s correlation coefficient (R) and p value of the trend lines shown in green. Each point represents a biological sample. **G-I,** Receiver Operating Characteristic (ROC) analysis of TIMP1 for the discrimination of Stroke/Control individuals and LE/OE individuals, respectively, with the Area Under the ROC Curve (AUC) shown in green. The black dots represent points on the ROC curve to gain optimal discriminative measures. **H-J,** ROC analysis of LGALS3 for the discrimination of Stroke/Control individuals and LE/OE individuals, respectively. The black dots represent points on the ROC curve to gain optimal discriminative measures.

Linear regression analysis was performed to rule out the effect of age on candidate protein levels. Correlations between TIMP1, LGALS3 levels and age using scatter plots depicted by trend lines shown as red dashed lines in Figure 5E and Figure 5F, separately. The Pearson’s correlation coefficient (R) values, along with their corresponding *p*-values, were displayed in green text on each trend line. All R values were less than 0.3 with *p*-values greater than 0.05, indicating no significant linear relationship between changes in TIMP1, LGALS3 levels and age.

Furthermore, Receiver Operating Characteristic (ROC) analysis were addressed to evaluate the discriminatory sensitivity and specificity between individuals with Stroke and Control. This step, depicted in green, represents optimal discriminative measures indicated by black dots on the ROC curve. Notably, TIMP1 (AUC = 0.904) and LGALS3 (AUC = 0.995) exhibited superior sensitivity (Figure 5G-H). We further examined the discriminatory sensitivity towards distinguishing LE/OE individuals, yielding AUC values of TIMP1 (0.873) and LGALS3 (0.794) more than 0.75 (Figure 5I-J), which suggested reasonable sensitivity and accuracy. These findings highlight that both TIMP1 and LGALS3 hold promise as reliable and practical biomarkers for predicting ischemic stroke rehabilitation.

### 3.3 Develop a combined prognosis panel for stroke rehabilitation

To evaluate clinical-based indicators for risk prediction in the subacute recovery of ischemic stroke, we collected multiple clinical data from 15 patients with LE and 11 patients with OE, including ADL assessment, age, grip strength, blood routine, urine routine, eight items of liver function, six items of blood lipids, six items of electrolytes, and 6 items of kidney function. In this step of the analysis, ADL recovery of the LE (Longshi Scale from Bedridden group to Bedridden group, BI from 23.67 to 25.67) and OE (Longshi Scale from Bedridden group to Domestic group, BI range from 30.91 to 55.45) groups presented noticeable difference (Table 2 and Figure S2A-B). Clinical test index data revealed significant differences by *t*-test in 9 clinical indicators such as hemoglobin (HGB), erythrocyte count (RBC), hematocrit, coefficient of variation of red blood cell distribution width (RDW-CV), low-density lipoprotein cholesterol (LDL-c), standard deviation of red blood cell distribution width (RDW-SD), uric acid (UA), total cholesterol (Cho) and albumin (ALB) as shown in Figure 6A and Table S8. Meanwhile, ROC analysis revealed that the area under curve (AUC) of 14 indicators exceeded 0.7, indicating excellent sensitivity and specificity in discriminating between LE/OE individuals. Among them, 9 out of the total 14 indicators were depicted in Figure 6B, exhibiting consistency with the indicators derived from the *t*-test analysis. Additionally, Figure S2C illustrated the AUC values for the remaining 5 indicators (age, lymphocyte count, lymphocyte ratio, Mean erythrocyte hemoglobin concentration, viscose silk). The AUC results in the graph indicate that a yellow background represents a positive correlation with good prognosis, while a red background represents a negative correlation. Data indicated that higher levels of HGB, RBCs, hematocrit, UA, and ALB were positively associated with good prognosis, while higher RDW-CV LDL-c RDW-SD, and Cho levels were negatively associated with good prognosis. The *t*-test and ROC analysis conducted above had demonstrated the reliability of the data quality in the enrolled patients. Consequently, these 9 clinical-based indicators were screened out for variable screening and modeling by machine learning to accurately predict prognosis of stroke recovery.

**Fig6.**
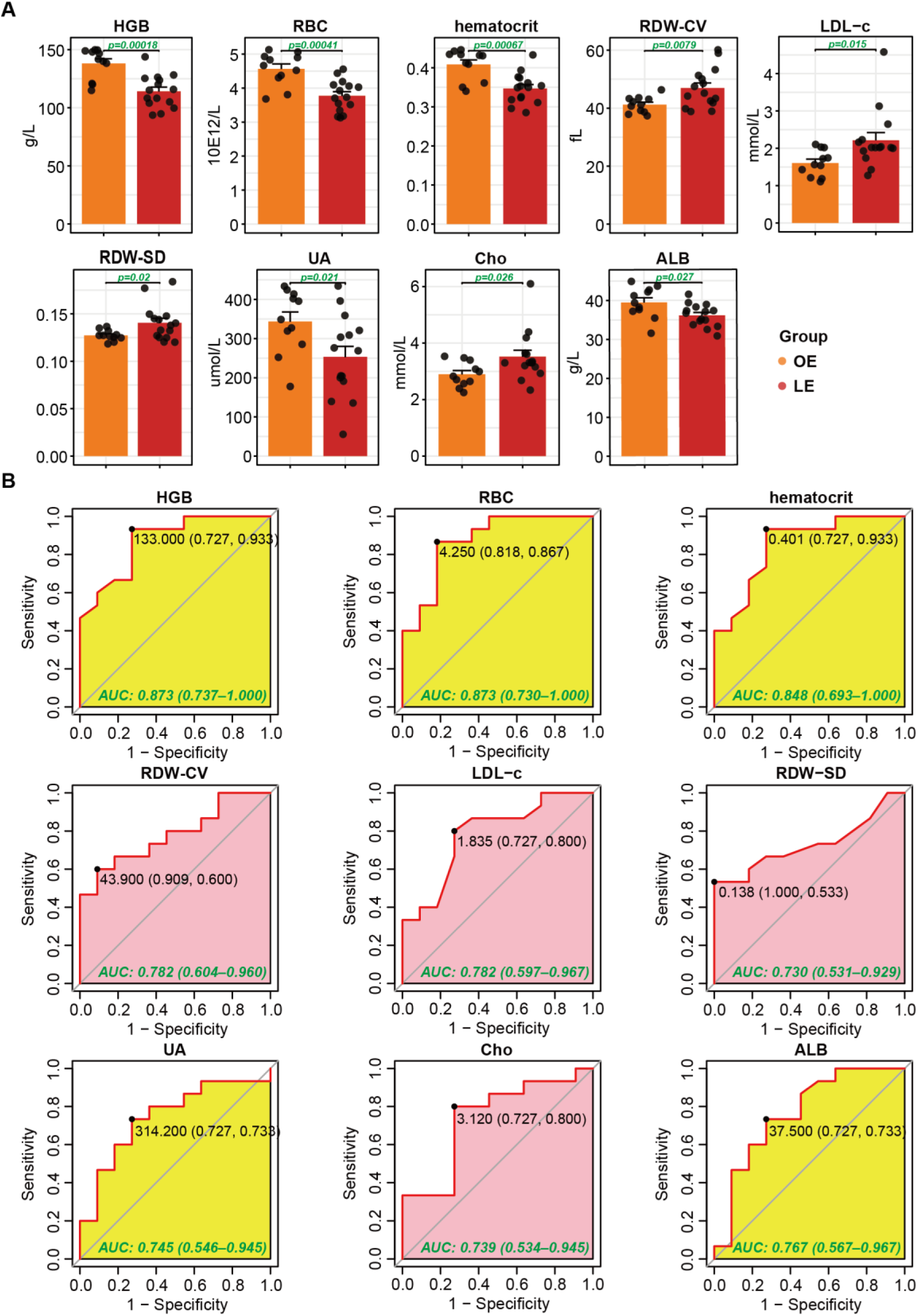
Analysis of clinical indicators of recruited clinical subjects with ischemic stroke. **A,** Histograms of the clinical indicator expression levels in the LE (n=15) and OE (n=11). The showed are the statistically significant indicators those with significant differences according to *p*-value from smallest to largest from left to right, and from up to down, and those without significant differences are shown in Supplementary Figure 2 C. Each point represents a biological sample. **B,** Receiver Operating Characteristic (ROC) analysis of indicators in A for the discrimination of LE/OE individuals, respectively, with the Area Under the ROC Curve (AUC) shown in green. The black dots represent points on the ROC curve to gain optimal discriminative measures. The yellow background represents a positive correlation with good prognosis, and the red background represents a negative correlation. **Abbreviations:** HGB, hemoglobin, RBC, erythrocyte count; RDW-CV, SD of erythrocyte volume; LDL-c, low-density lipoprotein cholesterol; RDW-SD, blood red cell volume distribution width; UA, uric acid; Cho, total cholesterol; ALB, albumin.

**Table 2.** ADL assessment of enrolled subjects for ELISA validation.

| Characteristic | Health Controls<br>(HC) | Ischemic Stroke (IS) |  |
| --- | --- | --- | --- |
|  |  | Obvious effects (OE) | Little effects (LE) |
| Gender (male) | 18 | 11 | 15 |
| Age (year) | 67.00 ± 2.099 | 63.27 ± 2.711 | 69.80 ± 2.322 |
| Diagnosis | Normal | Ischemic stroke | Ischemic stroke |
| ADL assessment of Barthel Index at admission (at discharge) | NA | 30.91 ± 3.149<br>(55.45 ± 6.923) ** | 23.67 ± 4.125<br>(25.67 ± 4.165) |
| ADL assessment of Longshi Scale at admission (at discharge) | NA | Bedridden group<br>(Domestic group) | Bedridden group<br>(Bedridden group) |

1 "\*\*\*" represents statistical significance at $p < 0.01$

In this study, we aimed to develop a combined prognostic model based on inflammation-derived biomarkers and filtered clinical-indicators for distinguishing subacute prognostication outcomes among patients with subacute ischemic stroke. The process and steps of establishing the prediction model using machine learning methods was illustrated in Figure 7A. The variable screening of machine learning was performed by LASSO analysis and Logistic regression, including those 9 clinical indicators (*p*-value < 0.05 in *t*-test and AUC > 0.7 in ROC) and 2 validated biomarkers (TIMP1 and LGALS3). Figures 7B-C present the coefficients, lambda correlation plot of LASSO filtered variables, and Lambda value plot of LASSO analysis. The absolute value of the index coefficient exceeding 0 in LASSO analysis and the *p*-value < 0.05 in logistic regression analysis served as the screening criteria, ultimately 6 molecules (HGB, UA, LDL-c, ALB, TIMP1, and LGALS3) were selected (Table S9). Furthermore, machine learning using ten standard two-categorical variable algorithms applied for modeling in the above training data set (Table S8) and prediction of stroke recovery in another new test dataset (LE: n=15; OE: n =11, Table S10). In the new test dataset, *t*-test showed there was no difference in age between the LE (mean 70.87 ± 2.808) and OE (mean 60.09 ± 2.630) groups (Table 3), similar to the inclusion data for the training set (Figure 7D). While most of 10 machine learning algorithms in training datasets achieved a prediction efficiency of 100%, the random forest algorithm model demonstrated the highest prediction efficiency in new test datasets, reaching 80% (Figure 7E). The best model of the random forest exhibited an AUC of prediction efficiency at 100% for trainset data and 80% for testset data (Figure 7F).

**Fig7.**
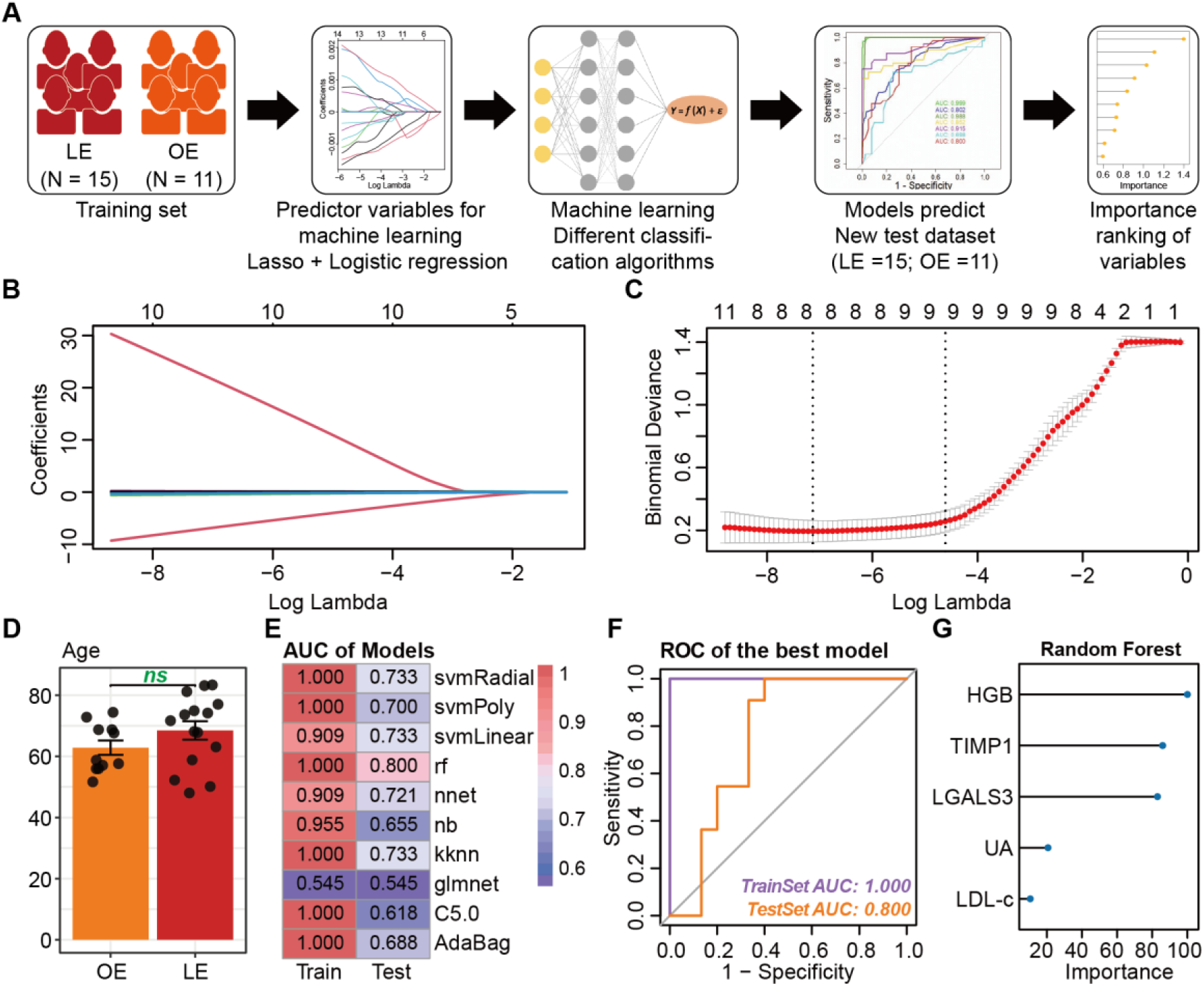
Screen biomolecules and clinical-based indicators to predict prognosis using machine learning modeling. **A,** Work flow chart of prognosis biomarker discovery with machine learning. **B,** Coefficients and lambda correlation plot of LASSO filtered variables. **C,** Lambda value plot of LASSO analysis. **D,** Age distribution statistics of clinical subjects recruited to the test dataset. Each point represents an individual. **E,** A total of 10 prediction models with 10 different machine learning classification algorithms for distinguishing dichotomous variables and the calculated discriminatory AUCs of each model across the training datasets (Train) and the blinded testing datasets (Test). Machine learning algorithms including Support Vector Machine (svmRatial, svmPloy, and svmLinear), Artificial Neural and Network (nnet), Logistic Regression (glmnet), Naive Bayesian Classifier (nb), K-Nearest Neighbor (kknn), Decision Tree Algorithm (C5.0, AdaBag, and Random Forest). **F,** ROC plots of the best-performing model in panel E, and the prognosis accuracy on the corresponding datasets were shown in the lower right corner. **G,** The top 5 feature molecules in the best predictive model (“random forest” on title) in panel F based on the variable importance ranking.

**Table 3.** ADL assessment of enrolled patients for test dataset.

| Characteristic | Ischemic Stroke (IS) |  |
| --- | --- | --- |
|  | Obvious effects (OE) | Little effects (LE) |
| Gender (male) | 11 | 15 |
| Age (year) | 60.09 ± 2.630 | 70.87 ± 2.808 |
| Diagnosis | Ischemic stroke | Ischemic stroke |
| ADL assessment of Barthel Index at admission (at discharge) | 32.73 ± 4.737<br>(57.27.45 ± 4.879) ** | 22.33 ± 3.268<br>(23.00 ± 3.229) |
| ADL assessment of Longshi Scale at admission (at discharge) | Bedridden group<br>(Domestic group) | Bedridden group<br>(Bedridden group) |

"\*\*\*" represents statistical significance at $p < 0.01$

An important advantage of the random forest model is that it can rank the importance of predictive features, which has important implications for clinical management. Except for ALB without any contribution value in the model with random forest, the importance of 5 indicators (HGB, TIMP1, LGALS3, UA, LDL-c) related to inflammatory response were visualized in Figure 7G. This study confirmed that central immune regulation might be crucial in stroke rehabilitation outcomes.

## 4. Discussion

We developed a machine learning-based model to predict functional rehabilitation of subacute ischemic stroke accurately. This interdisciplinary study, involving clinical and animal samples, provided robust evidence that a combined prediction model of incorporating serum biomarkers (TIMP1 and LGALS3) and clinical-based indices (HGB, UA, LDL-c), which primarily encompassed the pathophysiology of neuroinflammation, anemia, and antioxidation, might help to differentiation disease severity of recovery. Furthermore, a random forest of machine learning has demonstrated the combined model’s sensitivity and reliability in predicting ADL outcomes of post-stroke patients. Further extensive research is required to elucidate the function and underlying mechanisms by which the unbalanced neuroinflammatory response contributes to elevated TIMP1 and LGALS3 during stroke recovery.

The improvement of activity disorder from bedridden to domestic must result from the cooperation of various somatic functions and shows significant differences in biochemical indicators of nutritional status, neuroimmune, oxidative damage and repair, and innervation function, embodied in differences in protein function and levels. Although multiple traditional markers for the prediction of functional outcomes after ischemic stroke have been reported with reliable accuracy, there are few combined prediction models elaborated in the literature. In our study, after these multi-omics and multidimensional experiments explored and validated the practicable variables in serum, biological and clinical-based markers were first incorporated by a machine learning algorithm to accurately predict the outcome of subacute ischemic stroke rehabilitation, grouped subjects with the novel and convenient Longshi scale for monitoring disability stratification of ADL in post-stroke. The grouping of LE (recovery from bedridden to bedridden) and OE (recovery from bedridden to domestic) according to the assessment of ADL at admission and discharge by the Longshi scale was equivalently consistent with the grouping by BI. The Longshi scale may be an alternative grouping method for biological and clinical research analysis on optimizing biomarker performance.

Blood biomarkers have the potential to reflect underlying molecular and cellular processes, thereby facilitating the development of effective therapeutic strategies and improving rehabilitation outcomes. Functional rehabilitation after stroke events are highly dependent upon restitution, substitution, and compensation of neural network connectivity^16^; reliable predictors of stroke rehabilitation should play a crucial role in the process of injury and repair of NVU. Dead and dying cells or substances released by ischemic penumbra stimulate the production of inflammatory responses, mainly in microglia. Alternative immune activation is associated with decreased inflammation, decreased neurological damage, and increased NVU repair. Though advanced blood omics have benefited from the discovery of many new inflammation-related molecules, it is difficult to trace the central pathological changes directly in humans. Therefore, validating biomarkers in animal and cell models thus emerges as a viable solution. Candidates screened and validated through different dimensions may provide more reliable predictive value, helping to guide the development of new interventions.

We performed LC−MS/MS-based proteomic analysis in stroke subjects and investigated how these phenotypes correlate with 3-month recovery from ischemic stroke measured with ADL assessment. In the study, we identified 194 DEPs in LE vs HC and 174 DEPs in OE vs HC. To lessen the possible candidates, we used the RNA sequence of the ischemic prefrontal cortex in mice with MCAO at day 3 and day 7, equivalent to the clinical subacute stage of stroke recovery, considering the possibility of false positives of differential proteins due to the limited of clinical sample size. As a result, we focused on the 27 overlaps between DEPs and DEGs. Protein interaction network and functional analysis pointed directly to 8 target molecules of TIMP1, LGALS3, VIM, TGFB1, MYH9, CSF1R, GSN, and PTPRC. The qPCR validation of MCAO mice and OGD/R heMEC/D3 cells showed an important increase of the *TIMP1, TGFB1,* and *LGALS3* expression after ischemia reperfusion. The elevation of TIMP1 and LGALS3 were confirmed in an enlarged clinical serum sample, while the level of TGFB1 was not significantly different in the subjects of stroke and healthy control. TIMP1^17, 18^ and LGALS3^19, 20^, as novel inflammation-related factors, have been proven to be positively correlated with the poor outcomes of stroke and atherosclerosis. Therefore, our study focused on exploring the value of TIMP1 and LGALS3 in predicting the prognosis of stroke rehabilitation. LGALS3 and TIMP1, as novel inflammatory factors, may play an essential role in microglia-related inflammation regulation and promote NVU injury repair.

TIMP1, one of the tissue inhibitors of metalloproteinase (TIMP) family, inhibits matrix metalloproteinases (MMPs), a vital protein in maintaining the homeostasis of extracellular matrix structure and function. Recent studies have revealed that TIMP-1 possesses MMP-independent functions, acting as an emerging multifunctional cytokine through binding to cell surface receptors in developing central nervous system diseases and tumors^21–23^. The change of TIMP1 is consistent with the survey conclusion that higher TIMP1 levels were associated with increased risk of mortality and major disability after acute ischemic stroke in clinical^17^, and the gene expression of *Timp-1* was upregulated in infarction of the MCAO model^24, 25^. Our study demonstrated that the correlation between changes in TIMP1 within 3 months post-stroke and stroke recovery scores remained significant even after adjusting for baseline stroke severity. The latest study on the neuroinflammatory regulatory effect of TIMP1 through receptor-mediated signaling on the protection of the blood-brain barrier, independent of MMP9^23^, may provide novel insights into the reparative mechanism of TIMP1 in stroke injury. In our study we verified that the change in serum TIMP1 level was consistent with the variation tendency in proteomics. ROC analysis showed the reliability and specificity of TIMP1 (AUC=0.904, 0.873) in distinguishing the stroke from the healthy and LE from OE groups separately. The result indicated TIMP1 could be a marker to differentiate different recovery effects.

Another vital biomarker, LGALS3 (Galectin-3), a beta-galactosidase binding protein involved in microglial activation, a novel inflammatory factor known for its role in intravascular inflammation, lipid endocytosis, macrophage activation, cellular activation, and proliferation^26^. Many studies have revealed that galectin-3 plays an important role as a diagnostic or prognostic biomarker for neurodegenerative disorders, certain types of heart disease, viral infection, autoimmune disease, and tumors^27–29^. Additionally, LGALS3 could serve as a novel marker for the prediction of stroke clinical prognosis, positively associated with poor functional outcome and an increased risk of mortality in stroke patients^30,31^. Our data on the ROC analysis of LGALS3 (AUC=0.794) in the differentiation of the stroke with poor and good prognosis were consistent with this result. In molecular mechanism research, increasing evidence supports that LGALS3 modulates microglial activation under neurodegeneration conditions^32^. Except for the report that LGALS3 inhibits progressive fibrosis by modulating inflammatory profibrotic cascades^33^, few studies have elucidated the molecular mechanism of LGALS3 in post-infarction. Moreover, we detected the co-localized expression of TIMP1 and Lgals3 with microglia marker protein Iba1 with immunofluorescence, respectively. The findings of our study demonstrate firstly that microglia expressed TIMP1 and Lgals3 significantly increased during post-ischemic repair. It may be interesting to study further the mechanism of TIMP1 and Lgals3 regulation of microglial activation in ischemic stroke recovery.

Some clinical test indicators are also significant predictors. The data obtained from our enrolled patients underwent ROC analysis and LASSO-filtered variables to identify valuable predictive indicators, which primarily encompassed the pathophysiology of neuroinflammation, anemia, and antioxidation during the subacute stage of stroke. HGB levels are the gold standard for anemia, commonly and associated with poor outcome function after a stroke^34, 35^. The World Health Organization defines *anemia* as a hemoglobin level of less than 130 g/L. The average HGB level of patients in the OE group was 136.32 g/L, whereas in the LE group, it was 119.73 g/L, indicating an anemic state which may contribute to a poor prognosis. Low LDL-c is associated with a reduced risk of cardiovascular events and outcomes^36^. Higher serum UA levels have been proved to be an independent predictor of poor outcomes^37^. Though it has been reported UA as an independent predictor in stroke prognosis remains controversial, the role of UA in ischemic stroke pathophysiology is inseparable from oxidative damage^37, 38^. Age is linked to the long-term outcome of post-stroke rehabilitation, and the older with a stroke will probably result in worse outcomes^39^. Our study showed no important differences in age between enrolled patients with good or poor prognosis. Additionally, we observed a weak association between age and prognostic indicators (p < 0.3). These provided evidence that the OE group with good prognosis attributed to intervention therapy regardless of age. Variables selected in in this study could effectively reflect the underlying pathological progression of stroke recovery, thereby ensuring the predictive model of stroke rehabilitation with high accuracy.

This study is subject to certain limitations. Firstly, using a relatively small clinical sample in proteomics analysis may result in fewer differential proteins and limit the scope of bioinformatics analysis. To address this limitation, we strictly adhered to specific inclusion criteria, including age range (50-80 years), gender (male), and duration of onset (1-2 months) to minimize objective individual differences. Additionally, transcriptome changes at different stages of mice with MCAO were performed to validate our findings. Furthermore, in the validation experiments using ELISA analysis, we expanded the sample size to include 18 cases in the healthy control group and 52 cases in the stroke group. Recruiting larger sample sizes for model construction could yield even better prediction results. Despite the potential controversy surrounding the small sample size used in this study, it is essential to emphasize that our ultimate biomarkers have been confirmed credible and valuable through two conventional biomarker selection methods and machine learning techniques.

## Conclusions

In conclusion, we reported using machine learning to develop a novel combination prognostic model of inflammation-derived biomarkers (TIMP1, LGALS3) and clinical-based biomarkers (HGB, UA, LDL-c) in predicting the rehabilitation of ischemic stroke. Our work raises the exciting possibility that monitoring changes in inflammatory protein in ischemic stroke recovery could be used to gauge the severity of stroke and used in complementary to clinical prognostic variables to function outcome during stroke recovery.

### Abbreviations

IS: ischemic stroke
LE: little effective recovery
OE: obvious effective recovery
HC: healthy control
ADL: activities of daily living
TIMP1: Tissue inhibitor of metalloproteinase 1
LGALS3: Galectin-3
MMPs: Matrix metalloproteinases
HGB: hemoglobin
LDL-c: low-density lipoprotein cholesterol
UA: uric acid
ROC: receiver operating characteristic
AUC: area under curve
NVU: neurovascular unit
ROS: reactive oxygen species
NO: nitric oxide
TNF-α: tumor necrosis factor α
MCAO: middle cerebral artery occlusion
LC-MS/MS: liquid chromatography-tandem mass spectrometry analysis
UHPLC: ultra high-performance liquid chromatography
PASEF: parallel accumulation serial fragmentation
FPR: false-positive rate
DEPs: differentially expressed proteins
PCA: principal component analysis
DEGs: differentially expressed genes
GO: gene ontology
TGFB1: Transforming Growth Factor beta 1
PTPRC: Protein Tyrosine Phosphatase Receptor Type C
VIM: Vimentin
MYH9: Myosin Heavy Chain 9
CSF1R: Colony stimulating factor 1 receptor
GSN: Gelsolin
TTC: triphenyl tetrazolium chloride
OGD/R: oxygen-glucose deprivation/ reoxygenation
PVN: paraventricular nucleus of hypothalamus
ECM: extracellular matrix
BBB: blood-brain barrier
PASEF: parallel cumulative serial fragmentation
DEPs: differentially expressed proteins
DEGs: differentially expressed genes
GO: Gene Ontology
ALB: Albmin
svm: Support Vector Machine
rf: Random Forest
nnet: Artificial Neural and Network
nb: naive bayesian classifier
kknn: K-Nearest Neighbor
glmnet: Logistic Regression
C5.0: 
SEM: standard error of the mean.

## Data Availability

There is no conflict of interest for all authors who share data and materials in this article.

## Acknowledgements and Funding Sources

This study was supported by funds from the Natural Science Funding of China (No.82272598 and No.81901470 to Jiao Luo, No. 82205065 to Mingchao Zhou), the Science, Technology and Innovation Commission of Shenzhen (JCYJ20220530150407015 to Yulong Wang, JCYJ20230807115123047 to Mingchao Zhou, JCYJ20210324135804012 to Jiao Luo), Natural Science Foundation of Guangdong Province, China (No. 2020A1515011203), the Postdoctoral Science Foundation of China (No. 2019M663100).

## Supplemental Materials

Table S1-S10 and Figure S1-S2

## Author Contributions

L.J. and C.Y. proposed the conception and design; L.J., C.Y. and Z.M.C. performed research; L.J., H.Y.Y, Z.L.Y., and M.Y.Q. contributed acquisition of clinical data; L.J., T.Q.Y., L.J.B., and G.J. contributed acquisition of animal and cell experimental data; L.J., X.P., C.C.Q., H.M.L., and Z.X.H. contributed interpretation of clinical data; L.J. and C.Y. drafted the manuscript and revised it critically for important intellectual content; W.Y.L. supervised the experiments and revised and finalized the approval of the version to be published.

---

**FigS1.**
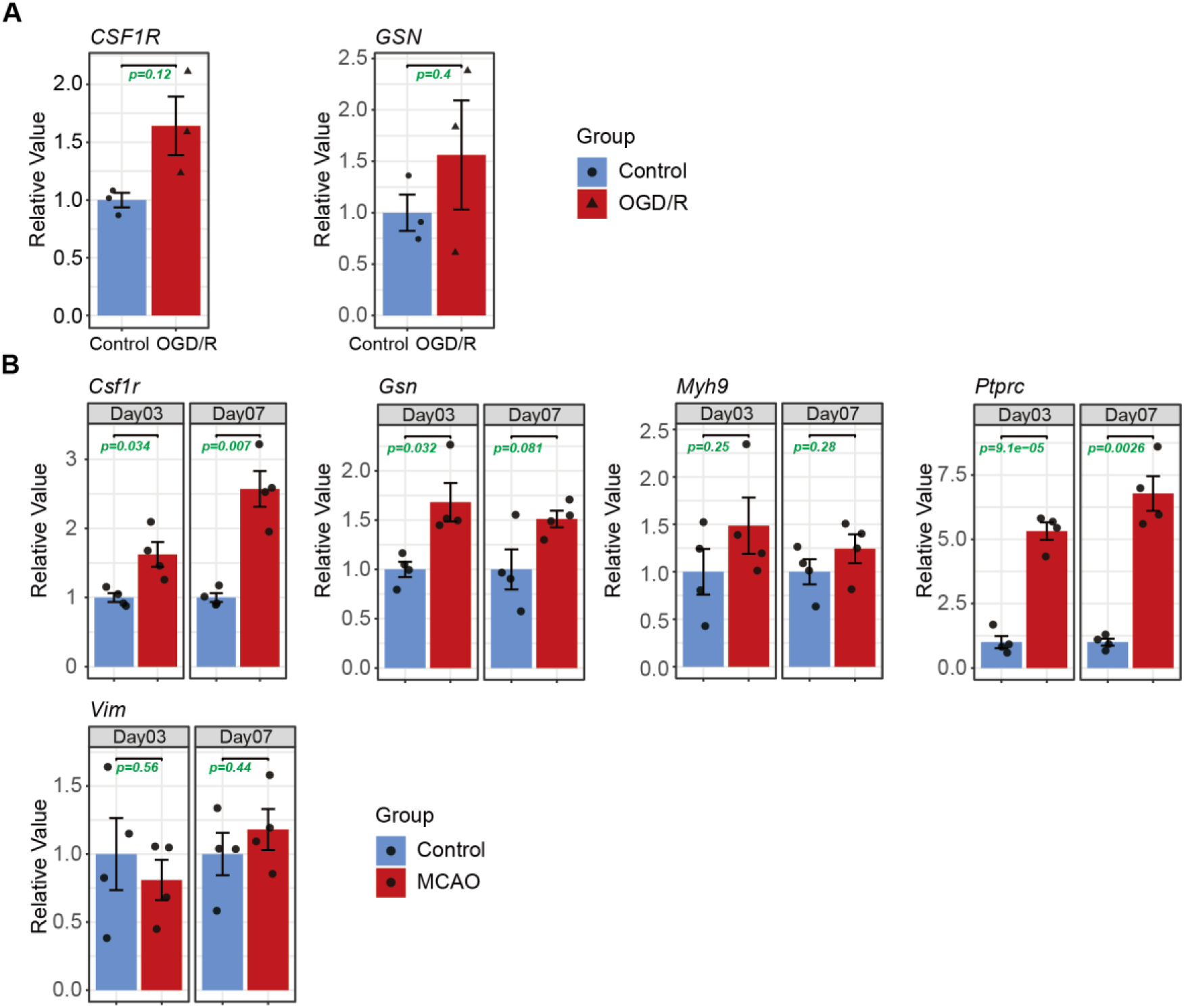
Supplementary figure corresponding to Figure 2. **A,** The histogram displays the qPCR expression levels of *CSF1R* and *GSN* mRNA in hcMEC/D3 cells after OGD/R. **B,** The histogram displays the qPCR expression levels of *Csf1r*, *Gsn, Myh9, Ptprc,* and *Vim* mRNA in ischemic cortex of mice with MCAO at day3 and day 7. Each data point represents a biological sample.

**FigS2.**
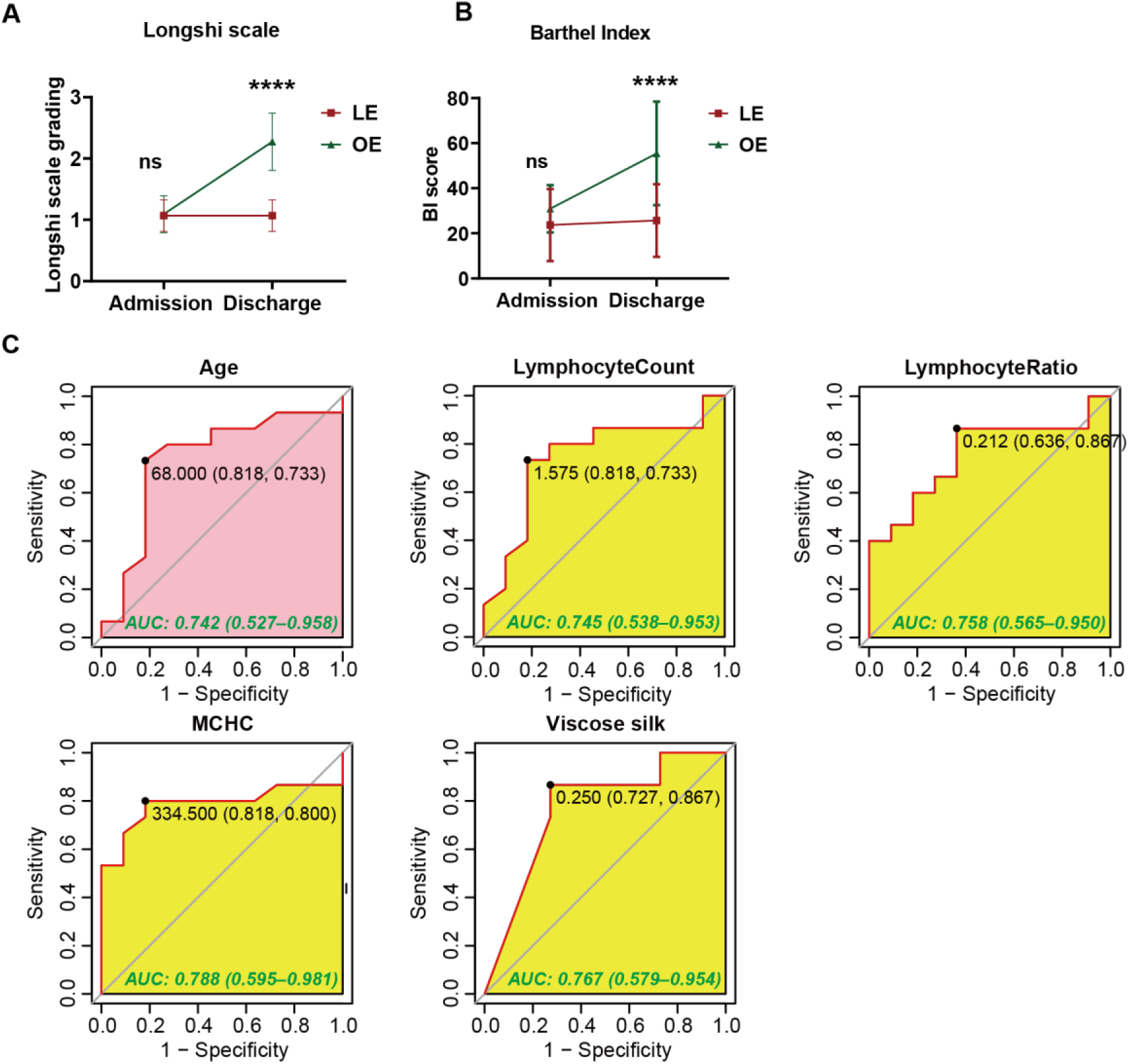
Supplementary figure corresponding to **Figure 5**. **A,** ADL assessment of the enrolled patients at admission and discharge by Longshi scale in the LE (n=15) and OE (n=11); **B,** ADL assessment of the enrolled patients at admission and discharge by Longshi scale, 1 = Bedridden group, 2 = Domestic group; **C,** ROC analysis of Age, LymphocyteCount, LymphocyteRatio, mean corpuscular hemoglobin concerntration (MCHC), and Viscose silk for the discrimination of LE/OE individuals, respectively.

